# Beyond Words: Non-verbal auditory cognitive impairments in Alzheimer’s Disease dementia

**DOI:** 10.1101/2024.09.02.24312935

**Authors:** Meher Lad, Charlotte Deasy, Christopher J Plack, John-Paul Taylor, Timothy D Griffiths

## Abstract

**Background:** Verbal speech-in-noise (SIN) measures are impaired in early Alzheimer’s disease (AD) but may be confounded by linguistic and cultural factors. We investigated whether non-verbal auditory memory could predict cognitive impairment in AD.

**Methods:** We evaluated 158 cognitively healthy individuals, 26 with mild cognitive impairment (MCI), and 28 with AD dementia using the Addenbrooke’s Cognitive Examination (ACE-III), pure-tone audiometry, verbal SIN tests, and non-verbal auditory memory tests for basic sound features. Group differences were assessed adjusting for age, sex, and education. Logistic regression and receiver operating characteristic (ROC) analyses compared model fit of verbal and non-verbal auditory variables.

**Results:** All auditory cognition measures were significantly associated with cognition. Non-verbal measures provided a better fit to diagnosis than verbal measures (AIC difference >10), although ROC analyses showed no significant differences between models.

**Conclusions:** Non-verbal auditory measures are effective measures in distinguishing between cognitively healthy, MCI, and AD dementia individuals.

## INTRODUCTION

Central hearing impairments can accompany cognitive impairment in Alzheimer’s disease (AD) and may precede the onset of dementia [1,2]. The underlying mechanisms linking central hearing and cognitive impairment in AD are not fully understood but some hypotheses suggest that it may relate to overlapping neuroanatomical substrates involved in speech-in-noise (SIN) hearing and neurodegeneration in AD [3]. This overlap implies that central auditory measures could serve as early indicators of neurodegenerative changes in AD, offering an avenue for screening and diagnostic assessments for AD dementia.

Verbal central auditory tasks have been commonly studied in people with AD dementia [4]. These tests commonly use digits, words or sentences as stimuli and performance is determined by a threshold measurement which signifies the ability of a person to correctly identify these on a background of artificial or natural sounds. In the digits-in-noise (DIN) test, a person is usually played three numbers on a background of white noise whereas in a sentence-in-babble test the stimulus consists of a short sentence on a background of multitalker babble. In cross-sectional studies, performance in both of these tasks reduces at a group-level from cognitively healthy to people with mild memory complaints to those with a dementia diagnosis [1]. Poor performance on both of these tests has also been linked to an increased risk of dementia, particularly AD, in independent studies [2,4].

Performance on these tests has been related to neurodegenerative biomarkers underlying the disease process [5]. One cross-sectional study showed an association between cerebrospinal fluid biomarkers of AD and performance on speech-in-noise perception tests. Poor auditory performance was correlated with high total-tau and phosphorylated-tau levels, after statistically adjusting for the effects of age, education, sex APOE4 status and pure-tone audiometry results.

Tau deposition in the brain is associated with cognitive impairment in AD and raised phosphorylated-tau in the cerebrospinal fluid is associated with a greater risk of AD dementia in the future [6,7]. Although this study was conducted in healthy individuals, this finding suggests a relationship between central hearing ability and neurobiological processes linked to an increased risk of AD dementia.

Speech-in-noise measures may, however, be confounded by linguistic proficiency, cultural background and educational attainment. These tests may therefore be less applicable to people from under-represented groups who are non-native speakers or those with low literacy levels. Such factors may lead to inaccurate measurements in these populations. Non-verbal measures of auditory cognition may be able to overcome these issues.

We used previously developed auditory cognitive tests to investigate how verbal and non-verbal metrics related to one another in ageing [8,9]. Auditory memory (the general ability to hold sound objects in mind) for basic sound features correlated with SIN perception ability as measured by sentence-in-babble thresholds. Verbal measures of auditory memory are also important determinants of SIN ability in older adults and those with hearing loss [10]. In previous work, we have found significant relationships with auditory memory for frequency and amplitude-modulation rate precision. We used a novel auditory paradigm which measured auditory memory as a continuous resource that is flexibly distributable to a sound object and provides a fine-grained measure of memory, rather than conventional discrete measures [11]. Further work established that including these measures in a linear model with educational attainment and verbal speech-in-noise measures allowed an accurate prediction of cognitive measures that are widely used as screening tools for dementia diagnosis [8].

In this study, we test the utility of non-verbal measures only in classifying a diagnosis of AD diagnosis. We hypothesised that non-verbal measures would be as effective as verbal SIN measures in distinguishing cognitive status among participants, including those of AD dementia. Therefore, the objectives of this study were: (1) to assess the utility of non-verbal auditory cognitive measures in distinguishing between different cognitive statuses associated with AD, and (2) to compare the effectiveness of non-verbal auditory measures with traditional verbal speech-in-noise tests in classifying cognitive impairment in AD.

We found that non-verbal auditory measures are as effective as verbal tests in distinguishing cognitive status among participants, including those with AD dementia. Non-verbal measures provided a better statistical fit for classifying cognitive status and were equally effective in discriminating between cognitively healthy individuals, those with mild cognitive impairment (MCI) and those with AD dementia. These results suggest that non-verbal auditory cognitive tests are valuable tools for detecting cognitive impairment associated with AD and may offer a more inclusive and generalisable approach for early detection and monitoring across diverse populations.

## METHODS

### Participants

Participants were recruited from the AudCog study, an ongoing observational study investigating auditory measures in cognitive health and disease. Inclusion criteria for cognitively healthy individuals included age between 50 and 85 years, normal cognitive screening scores (ACE-III ≥ 88) and no history of neurological or psychiatric disorders. Participants with MCI and AD dementia were also recruited and were diagnosed based on established clinical criteria by a neurologist or psychiatrist with specialist experience in dementia [12].

Participants were recruited from a variety of sources including local volunteer databases, the Join Dementia Research register and through the friends and family of patients from Memory Clinics for cognitively healthy participants. People with MCI and AD dementia were recruited through local memory and cognitive neurology clinics. In order to maximise diversity, equity and inclusion, we appealed to all individuals regardless of culture, race, gender or sexual orientation. A total of 212 participants were recruited: 158 cognitively healthy controls, 26 with MCI, and 28 with AD dementia. Demographic characteristics and auditory measurements are detailed in Table 1.

**Table 1.** Demographic and Auditory Characteristics.

|  | Cognitively Healthy<br>(n = 158) | Mild Cognitive<br>Impairment<br>(n=26) | Alzheimer's Disease<br>Dementia<br>(n=28) |
| --- | --- | --- | --- |
| *Demographic* |  |  |  |
| Age, mean (SD) | 67.0 (9.5) | 73.2 (6.9) | 75.2 (5.1) |
| Female, n (%) | 96 (62) | 16 (62) | 9 (38) |
| Advanced education, n (%) | 31 (20) | 4 (15) | 1 (4) |
| Hearing aid use, n (%) | 23 (15) | 4 (15) | 7 (29) |
| *Cognition* |  |  |  |
| ACE-III, mean/100 (SD) | 96 (3) | 84 (6) | 73 (9) |
| *Hearing measures* |  |  |  |
| Adjusted PTA threshold,<br>mean, dB HL (SD) | 44.8 (20) | 46.6 (17) | 40.6 (16) |
| DIN threshold, dB (SD) | -8.9 (4.1) | -7.5 (3.6) | -4.2 (2.9) |
| SIB threshold, dB (SD) | -6.9 (3.4) | -3.4 (6.9) | -0.5 (7.9) |
| AuMF, au (SD) | 0.29 (0.2) | 0.13 (0.1) | 0.09 (0.1) |
| AuMA, au (SD) | 0.08 (0.03) | 0.05 (0.03) | 0.04 (0.02) |

Abbreviations: ACE-III, Addenbrooke's Cognitive Examination (3rd Edition); au, arbitrary units; AuMA, Auditory Memory for Amplitude Modulation rate; AuMF, Auditory Memory for Frequency; DIN, Digits-in- Noise test; SIB, Speech-in-Babble test; PTA, Pure Tone Audiometry.

### Study Design

This study had a cross-sectional observational design. In-person written consent was obtained from all participants. All participants had one visit to the Auditory Laboratory at Newcastle University or a home visit was conducted to the participant’s home. After the consenting process, participants underwent a pure-tone audiometric test followed by auditory cognitive assessment. Finally, the cognitive screening test (described below) was administered. The recruitment period occurred from March 2022 to March 2024. Ethical approval was obtained from the Oxford C NHS Research Ethics Committee (21/SC/0139) and written informed consent was obtained from all participants. The study was performed in accordance with the ethical standards as laid down in the 1964 Declaration of Helsinki and its later amendments.

### Auditory Assessments

Pure tone audiometry (PTA) testing was performed on both ears from 250 Hz to 8 kHz at octave intervals for air conduction using an Interacoustics AD629 Diagnostic Audiometer and RadioEar DD450 circumaural headphones according to British Society of Audiology testing guidelines [13]. Pure tones were manually presented as short bursts twice starting at 30 dB HL then increased in 5 dB HL increments until comfortably audible if necessary. Then 5 dB HL reductions were made until the tone was not audible. This process was repeated twice, and the lowest audible volume was chosen as the value for a particular frequency. If maximum amplification at 100 dB HL could not be perceived, then this was used as the ceiling value at a particular frequency. The overall mean of high frequency values between 4 and 8 kHz for the best ear was taken as the threshold value for an individual for further analysis. This value was chosen as high-frequency thresholds are suspected to deteriorate first in age-related hearing loss and previous research from our group has suggested that PTA thresholds in this range correlate with speech-in-noise difficulties [14,15].

People wore Sennheiser Momentum 4 over-ear headphones for central auditory tests. Central hearing tests included verbal speech-in-noise perception tests using digits (Digits-in-Noise, DIN) and sentences (Speech-in-Babble, SIB). In these tests, the level difference between speech and background (white noise for DIN and 16-talker babble for SIB) is varied adaptively until a signal-to-masker ratio (in dB) at speech recognition threshold is obtained for each individual [8]. Participants had two practice trials at the beginning of the task to familiarise themselves with the stimuli at an SNR of 10 dB. An adaptive 1-up, 1-down psychophysical paradigm was implemented whereby a correct response resulted in the SNR being reduced and an incorrect one caused the SNR to increase. The starting SNR was 0 dB and the step sizes decreased from 5 to 2 dB after 3 reversals, which then reduced to 0.5 dB after 3 more reversals. The run terminated after 10 reversals and the SNR at the last 5 reversals was averaged to calculate the DIN threshold for each participant. Lower SNR values indicate a better performance. For the DIN task, select the digits they had heard from a keypad shown on the screen. For the SIB task, target sentences had the form 〈name〉〈verb〉 number〉 〈adjective〉 〈noun〉(e.g. “Alan gives four pretty flowers”) and participants had to click on the correct word from a list of five columns (10 options for each word) shown on the screen with the same structure.

Non-verbal auditory tests measure Auditory Memory (AuM) precision for basic sound features over several seconds. These tests are usually self-directed and take approximately 5-10 minutes. This tests short-term memory performance for two basic sound features: frequency of pure-tone and temporal fluctuation (modulation rate of amplitude-modulated white noise) [9]. A one-second tone or AM modulated white noise stimulus is presented to a participant after which they are asked to ‘find’ the sound on a horizontal scale on a computer screen. Participants move a mouse and click on the line to produce a sound at that location. They can make as many clicks as they want with no set time limit. After they were satisfied with their choice, they would advance to the next trial by pressing the ‘Enter’ key on a keyboard. Frequencies that determined the pure-tone sounds were chosen from a uniform distribution between 440 and 880 Hz and AM rates for the white noise stimulus were 5–20 Hz with a sinusoidal function used to apply this modulation. Hanning windows were applied to all synthetic sounds to avoid clicks and the beginning and end of the stimuli. The task consisted of 32 trials with the frequency and AM rate matching trials being interleaved. Participants had a short break after 16 trials. A Gaussian function was used to estimate the standard deviation of the errors in each trial across the whole experiment and the inverse of this value, the precision, was used for further analysis. Thus, one obtains precision scores for frequency AuM (AuMF) and AM rate AuM (AuMA). Studies in vision have found that this measure better reflects the memory resource a participant can allocate in a given task [16]. Participants had two practice trials with each stimulus (2 for frequency and 2 for AM rate AuM) at the beginning of the task to familiarise themselves with the stimuli.

### Cognitive Assessment

All participants underwent a cognitive screening test using the Addenbrooke’s Cognitive Examination 3rd Edition (ACE-III). A score of 88 is commonly used to delineate cognitive impairment [17]. The ACE-III demonstrated variable sensitivity (ranging from 82% to 97% for dementia at thresholds of 82 and 88) and specificity (ranging from 4% to 77% for dementia) across different thresholds and patient populations, with more variability observed in specificity.

### Statistical analysis

All variable data were checked for normality by visually examining their distributions. AuM values were log-transformed to achieve normality. Mean and standard deviation (SD) values were calculated using absolute values in order to facilitate comparison between groups using an

Analysis of Variance (ANOVA). To assess group differences across cognitive and auditory measures, one-way ANOVA was performed for each outcome variable: adjusted ACE-III total score, DIN, SIB, AuMF and AuMA z-scores. Each ANOVA tested for differences across three cognitive groups: Cognitively Healthy (CN), MCI, and AD Dementia. Post-hoc analyses using Tukey’s HSD test were conducted to explore pairwise group differences where applicable. All analyses controlled for age, sex at birth, and educational status, and significance was set at *p* < 0.05. To aid analysis using linear models, these values were standardised using z-scores.

Multinomial logistic regression models were used to assess the association between the non-verbal auditory measures and clinical diagnosis, using age, sex and educational attainment as regressors of no interest. In order to compare this model to one created using verbal measures, we used Bayesian Model Comparison: Model 1 with the DIN and SIB tests and Model 2 with AuMF and AuMA. We used the Akaike Information Criterion (AIC) to define the best model. A lower AIC with a difference of 2 from the other model was used to define an informative model with parsimonious degrees of freedom. The models were also evaluated and compared (using bootstrapped confidence intervals) for classification accuracy for diagnosis using Receiver Operator Characteristic (ROC) curves.

All statistical analyses were conducted using Python programming language within Jupyter notebooks and Visual Studio Code environments. We utilised several Python libraries to perform data processing and statistical modelling. Specifically, we employed statsmodels for implementing multinomial logistic regression models and assessing model parameters. The patsy library was used for creating design matrices necessary for the regression analyses. To evaluate multicollinearity among predictors, we calculated the Variance Inflation Factor using functions from statsmodels.stats.outliers_influence. For data preprocessing, including standardisation and label binarization, we used functions from scikit-learn (sklearn), specifically the preprocessing module. The dataset was split into training and testing subsets using train_test_split from sklearn.model_selection. Multinomial logistic regression models were implemented using OneVsRestClassifier and LogisticRegression from sklearn.linear_model. Receiver Operating Characteristic (ROC) curves were generated, and the Area Under the Curve (AUC) values were calculated using functions from sklearn.metrics. This allowed us to assess the discriminative power of the verbal and non-verbal auditory measures in classifying cognitive status.

## RESULTS

As shown in Table 1, cognitively healthy people were younger and better educated than those with mild cognitive impairment or AD dementia and therefore these measures were used as regressors of no interest along with sex at birth. Cognitive scores on the ACE-III also decreased from cognitively normal to mild cognitively impaired people to people with a diagnosis of AD dementia. Adjusted pure-tone audiometric thresholds did not differ between groups. The unadjusted average audiograms for each group are shown in Figure 1.

**Figure 1.**
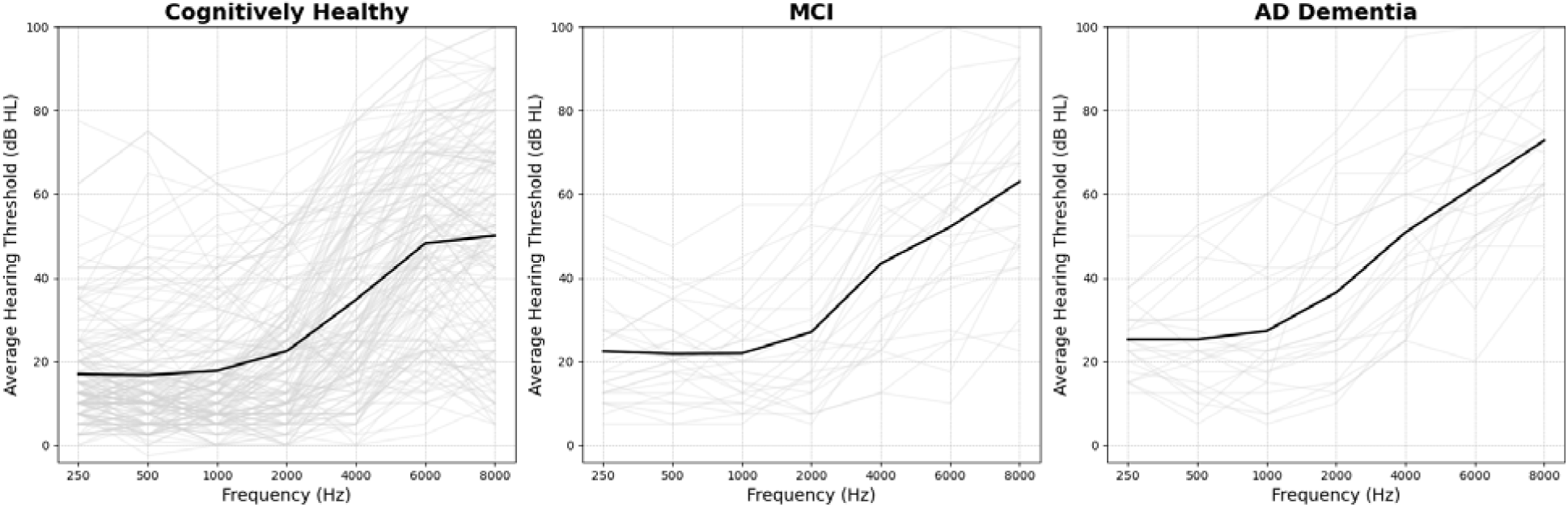
Pure-tone audiogram results for cognitively healthy, Mild Cognitive Impairment (MCI) and Alzheimer’s Disease (AD) dementia groups. The figure presents the unadjusted averaged hearing thresholds (dB HL) across frequencies (250–8000 Hz) for three cognitive groups: Cognitively Healthy, MCI, and AD Dementia. Individual participants’ audiograms are shown as translucent grey lines, while the group average for each frequency is represented by a solid black line. Hearing thresholds are averaged between the left and right ears for each participant. This figure illustrates a general trend of increasing hearing impairment with higher frequencies across all groups.

Statistical analysis revealed significant differences in cognitive and auditory performance across groups (Figure 2). ANOVA results showed that ACE-III total scores differed significantly between groups (F(2, 203) = 325.47, p < 0.0001), with lower scores in the MCI and AD groups compared to cognitively healthy participants. Similarly, significant group differences were observed in auditory measures: DIN (F(2, 203) = 9.24, p < 0.001), SIB (F(2, 203) = 45.63, p < 0.001), AuMF (F(2, 203) = 14.57, p < 0.0001), and AuMA (F(2, 203) = 28.43, p < 0.001). Figure 3 further highlights significant correlations between the ACE-III total score and auditory measures. Strong correlations were observed for both verbal (SIB; r = 0.74, p < 0.001) and non-verbal measures (AuMA; r = 0.58, p < 0.001), indicating that auditory processing, both verbal and non-verbal, declines in parallel with cognitive impairment.

**Figure 2.**
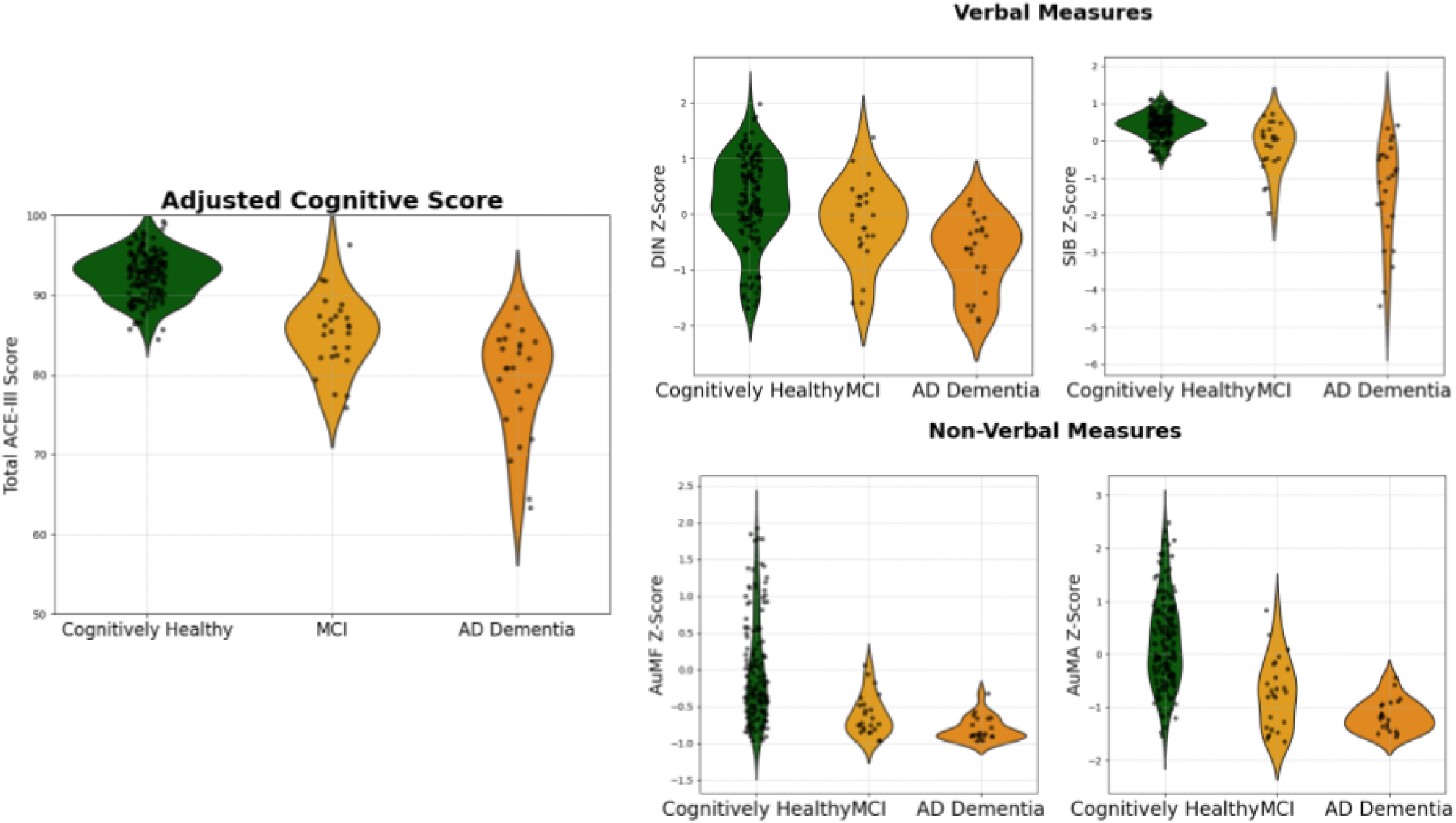
Cognitive and auditory measure distributions by participant group. The left panel shows the adjusted ACE-III total scores for three cognitive groups: Cognitively Healthy (CN), Mild Cognitive Impairment (MCI), and Alzheimer’s Dementia (AD). The right panel is split into two sections: Verbal Measures (top) and Non-Verbal Measures (bottom). The Verbal Measures display violin plots for the Digits in Noise (DIN) and Speech in Babble (SIB) z-scores, while the Non-Verbal Measures show distributions for Auditory Memory for Frequency (AuMF) and Auditory Memory for Amplitude Modulation (AuMA) z-scores. Each plot visualises the distribution of individual scores within each cognitive group, with individual data points overlaid in black.

**Figure 3.**
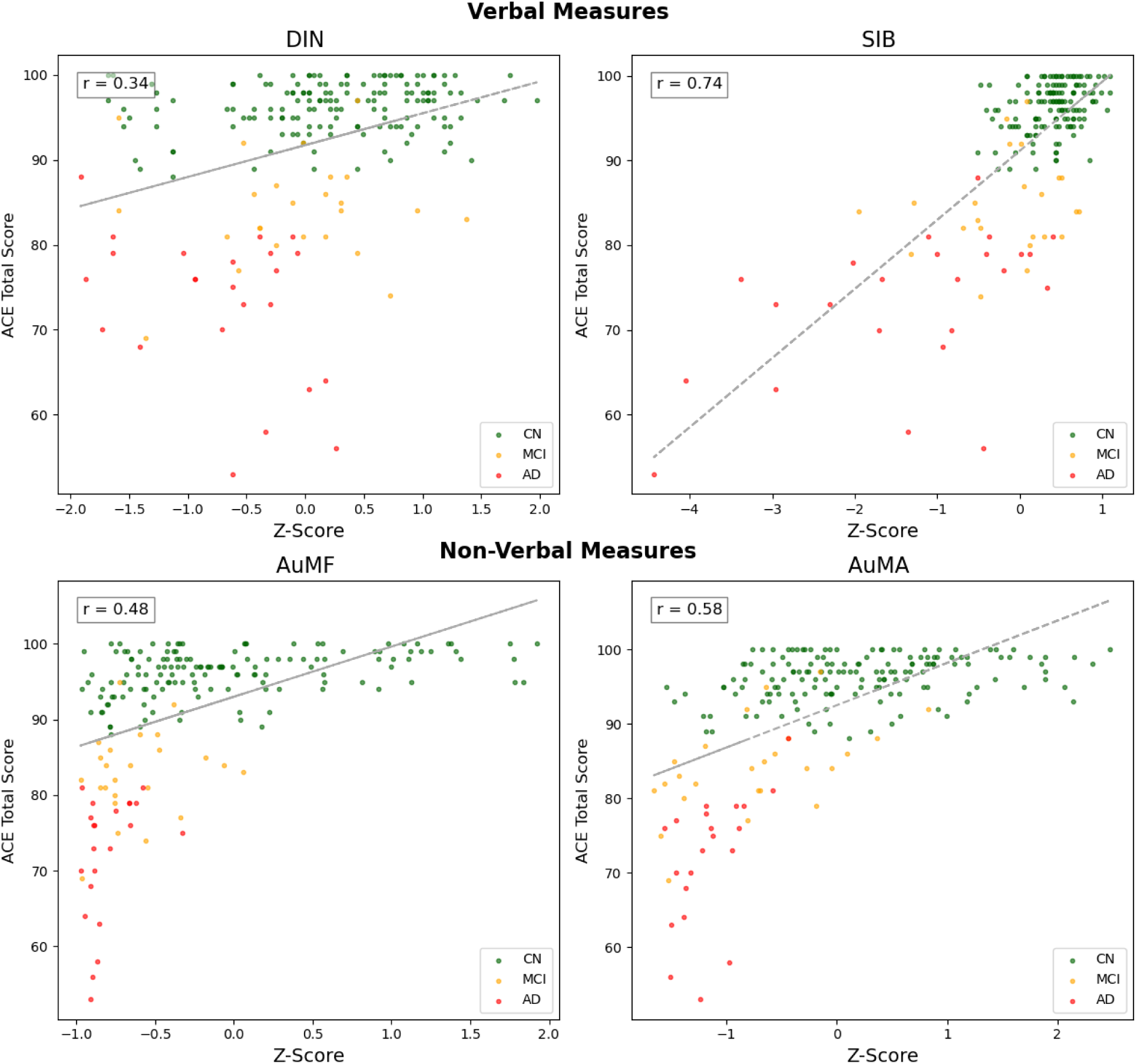
Correlations between cognitive and auditory measures and ACE-III total score. This figure shows scatterplots of four cognitive and auditory measures, grouped by cognitive status (Cognitively Healthy (CN), Mild Cognitive Impairment (MCI), Alzheimer’s Disease (AD) dementia) and their relationship with the ACE-III total score. The upper row (Verbal Measures) presents the associations for the Digits in Noise (DIN) and Speech in Babble (SIB) z-scores, while the lower row (Non-Verbal Measures) presents the associations for Auditory Memory for Frequency (AuMF) and Auditory Memory for Amplitude Modulation (AuMA) z-scores. Trendlines (dark grey) highlight the correlations, with corresponding r-values displayed in each subplot. All correlations were statistically significant with p < 0.001.

**Figure 4.**
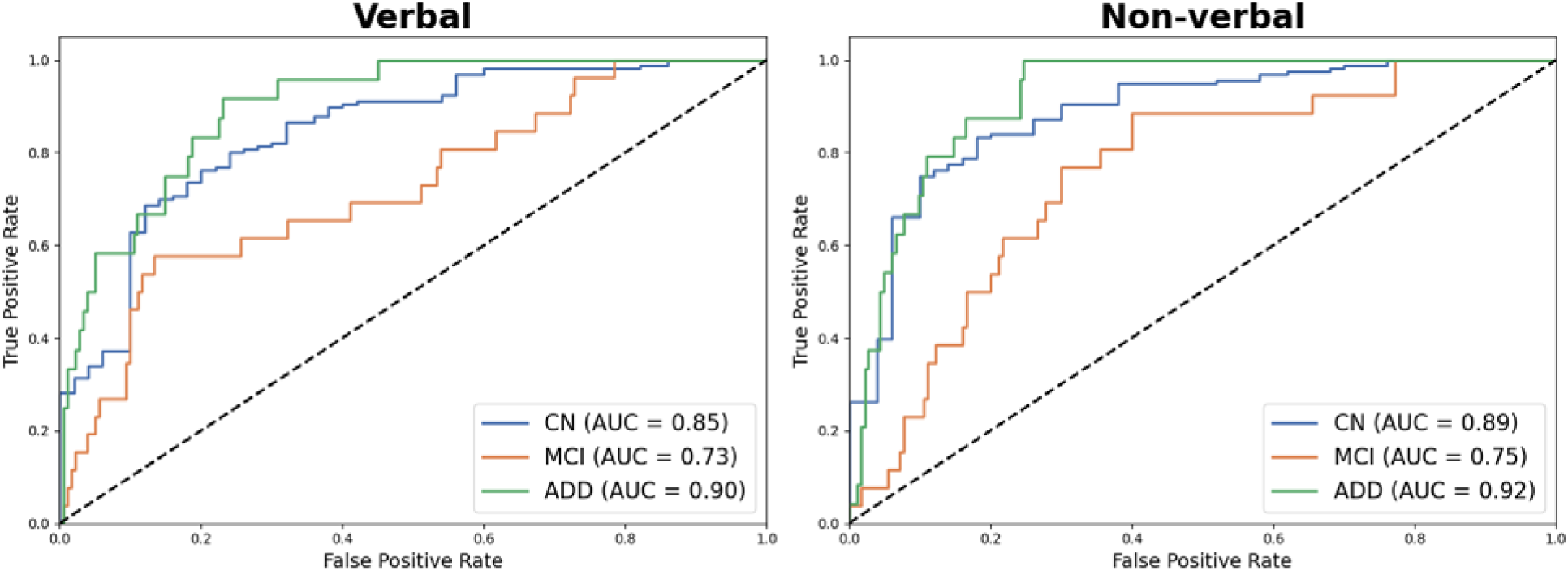
Receiver Operating Characteristic (ROC) Curves for Verbal and Non-Verbal Auditory Measures in Discriminating Cognitive Status. The figure displays the ROC curves for two multinomial logistic regression models used to differentiate between cognitively normal (CN), mild cognitive impairment (MCI) and Alzheimer’s disease dementia (ADD) groups based on auditory performance. The leftward image represents the model using verbal auditory measures, specifically the digits-in-noise performance and sentence-in-babble threshold. The rightward image shows the model using non-verbal auditory measures, including auditory memory for frequency and auditory memory for amplitude modulation rate. Each ROC curve corresponds to the classification of one cognitive status group (CN, MCI, ADD). The area under the curve (AUC) values are reported for each ROC curve, indicating the model’s ability to correctly classify people across cognitive statuses. Both verbal and non-verbal models demonstrate similar AUC values, suggesting that non-verbal auditory measures are as effective as verbal measures in associating with cognitive impairment.

We conducted multinomial logistic regression analyses to compare the effectiveness of verbal and non-verbal auditory measures in distinguishing between cognitively normal, mild cognitive impairment, and AD dementia groups. Model 1, which included verbal measures, explained 26% of the variance in cognitive status (Log-likelihood: -110.56, AIC: 247). Within this model, SIB had the largest effect on both mild cognitive impairment and AD dementia, indicating its strong association with cognitive impairment. Model 2 focused on non-verbal measures and demonstrated a better fit, explaining 32% of the variance (Log-likelihood: -100.99, AIC: 228). In this model, auditory memory for amplitude modulation rate had the largest effect size, significantly associated with both mild cognitively impaired people and people with AD dementia. The comparison of AIC values suggests that the non-verbal measures model (Model 2) provided a statistically better fit than the verbal measures model (Model 1).

To further assess the discriminative power of the verbal and non-verbal auditory measures, ROC curve analysis was performed, and the Area Under the Curve (AUC) values were calculated for each model (Figure 1). The verbal measures model produced AUCs of 0.85 for cognitively healthy, 0.78 for mild cognitively impaired people, and 0.82 for people with AD dementia, indicating a high level of accuracy in distinguishing between the cognitive groups. Similarly, the non-verbal measures model yielded AUCs of 0.83 for cognitively healthy, 0.79 for mild cognitively impaired people, and 0.80 for people with AD dementia, demonstrating that non-verbal measures were equally effective in classifying cognitive status. The comparative analysis of the ROC curves showed no significant difference in the AUC values between the verbal and non-verbal models (p > 0.05), suggesting that non-verbal auditory tests are as effective as verbal tests in identifying cognitive impairment.

## DISCUSSION

We demonstrate that simple non-verbal measures of auditory cognition are significantly associated with cognitive impairment in AD and can effectively discriminate between individuals with and without AD dementia. Auditory memory is linked to cognition across the AD spectrum and, when combined with demographic factors, can classify a clinical diagnosis. These findings lay the groundwork for future investigations into auditory memory as a potential measure of brain function, alongside fluid biomarkers that are sensitive and specific to AD, offering a more inclusive and accessible diagnostic tool.

Previous clinical observations in individuals with AD dementia have highlighted an aversion or heightened sensitivity to sound processing, particularly in noisy environments. However, detailed evaluations of these symptoms using non-verbal auditory paradigms are only now beginning to emerge [18]. Our results align with prior research showing impairments in auditory scene analysis and spatial processing when using complex sound stimuli [19,20]. Additionally, this study extends our understanding of non-verbal auditory cognition beyond individuals without dementia to those experiencing memory and cognitive difficulties due to AD dementia.

Auditory memory is a crucial cognitive ability that supports SIN perception, particularly as one ages [10,21]. Successful SIN perception requires the listener to attend to the correct auditory stream, track it and remember it over time despite interference from competing sounds. This process likely involves several cognitive factors. Previous research suggests that phonological auditory memory becomes increasingly important as individuals age and experience hearing loss [21]. While the underlying reasons for this are unclear, they may be related to the increased working memory resources required to comprehend sentences in noisy environments when perceptual systems are partially degraded.

The auditory memory precision measure, which is based on a continuous scale rather than conventional discrete phonological measures, has been shown to reflect the total memory ‘resource’ an individual can allocate to a given task [16]. The specific reasons for the associations between SIN perception and auditory memory for features such as frequency precision and amplitude modulation rates remain uncertain. This may relate to the evolutionary importance of humans tracking the source of a sound object e.g. voices, where frequency is particularly important and the natural modulations that occur in the envelope of speech sounds (which were similar to the rates used as stimuli in this study) [22–24].

The reasons for specific associations with auditory memory and cognition in AD have also not yet been thoroughly investigated. Some hypotheses propose that non-verbal measures of auditory memory link speech-in-noise perception ability and cognition in AD due to their neuroanatomical substrates [3]. Performing these tasks is known to activate medial temporal lobe structures, regions implicated in the early stages of AD pathology [25,26]. Degeneration of these structures could impair auditory memory and contribute to cognitive deficits detected by other screening tools. Additionally, damage to these areas, regardless of aetiology, can similarly affect performance in visual cognitive tasks [27,28]. It is also important to recognise that hearing function, age and brain neuropathology independently influence cognition and may collectively contribute to cognitive decline in AD [29].

This study benefits from incorporating both peripheral and detailed central auditory testing in individuals with and without dementia. Including such comprehensive measures is crucial, as a combination of peripheral and central auditory assessments may provide a more complete understanding of the neurobiological link between hearing and dementia. Previous studies relying solely on pure-tone audiograms or subjective measures have not consistently demonstrated relationships with the underlying neurobiological processes in AD [30,31]. This contrasts with some work using central measures [5]. Central auditory measures may also help clarify whether hearing aids can prevent or delay dementia. The lack of success in trials using hearing aids as an intervention may be due to their limited effectiveness in individuals with central auditory impairments, suggesting that hearing aids might be most beneficial for those without central auditory dysfunction [32].

Limitations of this study include the relatively small sample size of cognitively impaired individuals, the cross-sectional design and the lack of neurodegenerative biomarkers to characterise the participants. The progression of cognitive impairment in ageing is known to vary based on biomarker status and future work should incorporate these *in-vivo* markers to provide a more comprehensive understanding of the relationship between auditory cognition and AD progression [33]. While participants with normal cognitive function were younger and had higher education levels than other groups, these factors were statistically controlled for in the analyses. Future studies should aim to include more diverse populations, as these auditory measures are applicable across various cultural and linguistic backgrounds. Longitudinal research is also necessary to determine whether auditory cognition measures can predict the onset or progression of AD dementia, particularly in preclinical or mildly impaired individuals.

This work showcases simple non-verbal auditory stimuli which could potentially be used alongside clinical and neurodegenerative biomarkers for screening and monitoring cognitive impairment due to AD. These measures are potentially less susceptible to confounding factors such as linguistic and cultural differences, offering a broader applicability in diverse populations. Additionally, their suitability for use in remote or resource-limited settings enhances their practical utility [34]. Further work is necessary to see whether these non-verbal auditory tasks, like verbal SIN measures, can predict AD dementia, particularly in individuals with MCI [2].

Studying auditory memory alongside highly sensitive and specific fluid biomarkers of AD, such as phosphorylated-Tau, would explain whether poor auditory cognition is a harbinger of dementia [6]. Auditory cognition may also be associated with more general markers of neurodegeneration, such as cortical thinning, detectable through neuroimaging. Urgent work is required to investigate the relationship between auditory cognitive decline and these neuroimaging metrics [35].

In conclusion, this study demonstrates that non-verbal auditory memory tasks are strongly associated with cognitive impairment in Alzheimer’s disease and show potential as early indicators of cognitive decline. These tasks, particularly when combined with demographic factors,could help distinguish AD dementia from normal cognitive ageing. The findings suggest that non-verbal auditory measures tap into neural processes that are particularly vulnerable to early AD pathology, especially in medial temporal lobe structures. Further longitudinal studies incorporating auditory cognition, fluid biomarkers and neuroimaging are essential to clarify the role of auditory memory in predicting dementia and to explore whether these measures could enhance early detection or improve the monitoring of AD progression.

## Data Availability

All data produced in the present study are available upon reasonable request to the authors

## Acknowledgements

The authors would like to thank all the participants and their carers for participating in this research.

## Contributors

ML conceived the study. ML collected and analysed the data. ML drafted the initial manuscript. All authors contributed to revision and editing of the manuscript.

## References

[1] Gates GA, Anderson ML, Feeney MP, McCurry SM, Larson EB. Central Auditory Dysfunction in Older Persons With Memory Impairment or Alzheimer Dementia. Arch Otolaryngol Neck Surg 2008;134:771–7. 10.1001/archotol.134.7.771.

[2] Stevenson JS, Clifton L, Kuźma E, Littlejohns TJ. Speech-in-noise hearing impairment is associated with an increased risk of incident dementia in 82,039 UK Biobank participants. Alzheimers Dement J Alzheimers Assoc 2022;18:445–56. 10.1002/alz.12416.

[3] Griffiths TD, Lad M, Kumar S, Holmes E, McMurray B, Maguire EA, et al. How Can Hearing Loss Cause Dementia? Neuron 2020;108:401–12. 10.1016/j.neuron.2020.08.003.

[4] Mohammed A, Gibbons LE, Gates G, Anderson ML, McCurry SM, McCormick W, et al. Association of Performance on Dichotic Auditory Tests With Risk for Incident Dementia and Alzheimer Dementia. JAMA Otolaryngol--Head Neck Surg 2022;148:20–7. 10.1001/jamaoto.2021.2716.

[5] Tuwaig M, Savard M, Jutras B, Poirier J, Collins DL, Rosa-Neto P, et al. Deficit in Central Auditory Processing as a Biomarker of Pre-Clinical Alzheimer’s Disease. J Alzheimers Dis 2017;60:1589–600. 10.3233/JAD-170545.

[6] Palmqvist S, Tideman P, Cullen N, Zetterberg H, Blennow K, Alzheimer’s Disease Neuroimaging Initiative, et al. Prediction of future Alzheimer’s disease dementia using plasma phospho-tau combined with other accessible measures. Nat Med 2021;27:1034–42. 10.1038/s41591-021-01348-z.

[7] Bejanin A, Schonhaut DR, La Joie R, Kramer JH, Baker SL, Sosa N, et al. Tau pathology and neurodegeneration contribute to cognitive impairment in Alzheimer’s disease. Brain J Neurol 2017;140:3286–300. 10.1093/brain/awx243.

[8] Lad M, Taylor J-P, Griffiths TD. The contribution of short-term memory for sound features to speech-in-noise perception and cognition. Hear Res 2024;451:109081. 10.1016/j.heares.2024.109081.

[9] Lad M, Holmes E, Chu A, Griffiths TD. Speech-in-noise detection is related to auditory working memory precision for frequency. Sci Rep 2020;10:13997. 10.1038/s41598-020-70952-9.

[10] Akeroyd MA. Are individual differences in speech reception related to individual differences in cognitive ability? A survey of twenty experimental studies with normal and hearing-impaired adults. Int J Audiol 2008;47 Suppl 2:S53–71. 10.1080/14992020802301142.

[11] Joseph S, Teki S, Kumar S, Husain M, Griffiths TD. Resource allocation models of auditory working memory. Brain Res 2016;1640:183–92. 10.1016/j.brainres.2016.01.044.

[12] McKhann GM, Knopman DS, Chertkow H, Hyman BT, Jack CR, Kawas CH, et al. The diagnosis of dementia due to Alzheimer’s disease: recommendations from the National Institute on Aging-Alzheimer’s Association workgroups on diagnostic guidelines for Alzheimer’s disease. Alzheimers Dement J Alzheimers Assoc 2011;7:263–9. 10.1016/j.jalz.2011.03.005.

[13] British Society of Audiology. Pure-tone air-conduction and boneconduction threshold audiometry with and without masking 2018.

[14] Holmes E, Griffiths TD. ‘Normal’ hearing thresholds and fundamental auditory grouping processes predict difficulties with speech-in-noise perception. Sci Rep 2019;9:1–11. 10.1038/s41598-019-53353-5.

[15] Tl W, R C, L C, Dm N, Kj C. Changes in hearing thresholds over 10 years in older adults. J Am Acad Audiol 2008;19. 10.3766/jaaa.19.4.2.

[16] Ma WJ, Husain M, Bays PM. Changing concepts of working memory. Nat Neurosci 2014;17:347–56. 10.1038/nn.3655.

[17] Beishon LC, Batterham AP, Quinn TJ, Nelson CP, Panerai RB, Robinson T, et al. Addenbrooke’s Cognitive Examination III (ACELIII) and miniLACE for the detection of dementia and mild cognitive impairment. Cochrane Database Syst Rev 2019;2019:CD013282. 10.1002/14651858.CD013282.pub2.

[18] Fletcher PD, Downey LE, Golden HL, Clark CN, Slattery CF, Paterson RW, et al. Auditory hedonic phenotypes in dementia: A behavioural and neuroanatomical analysis. Cortex 2015;67:95–105. 10.1016/j.cortex.2015.03.021.

[19] Goll JC, Kim LG, Ridgway GR, Hailstone JC, Lehmann M, Buckley AH, et al. Impairments of auditory scene analysis in Alzheimer’s disease. Brain J Neurol 2012;135:190–200. 10.1093/brain/awr260.

[20] Golden HL, Nicholas JM, Yong KXX, Downey LE, Schott JM, Mummery CJ, et al. Auditory spatial processing in Alzheimer’s disease. Brain J Neurol 2015;138:189–202. 10.1093/brain/awu337.

[21] Füllgrabe C, Rosen S. On The (Un)importance of Working Memory in Speech-in-Noise Processing for Listeners with Normal Hearing Thresholds. Front Psychol 2016;7:1268. 10.3389/fpsyg.2016.01268.

[22] Assmann PF, Summerfield Q. The contribution of waveform interactions to the perception of concurrent vowels. J Acoust Soc Am 1994;95:471–84. 10.1121/1.408342.

[23] Oxenham AJ. Pitch Perception. J Neurosci 2012;32:13335–8. 10.1523/JNEUROSCI.3815-12.2012.

[24] Goswami U. Speech rhythm and language acquisition: an amplitude modulation phase hierarchy perspective. Ann N Y Acad Sci 2019;1453:67–78. 10.1111/nyas.14137.

[25] Kumar S, Joseph S, Gander PE, Barascud N, Halpern AR, Griffiths TD. A Brain System for Auditory Working Memory. J Neurosci Off J Soc Neurosci 2016;36:4492–505. 10.1523/JNEUROSCI.4341-14.2016.

[26] Kumar S, Gander PE, Berger JI, Billig AJ, Nourski KV, Oya H, et al. Oscillatory correlates of auditory working memory examined with human electrocorticography. Neuropsychologia 2020;150:107691. 10.1016/j.neuropsychologia.2020.107691.

[27] Borders AA, Ranganath C, Yonelinas AP. The hippocampus supports high-precision binding in visual working memory. Hippocampus 2022;32:217–30. 10.1002/hipo.23401.

[28] Zokaei N, Nour MM, Sillence A, Drew D, Adcock J, Stacey R, et al. Binding deficits in visual short-term memory in patients with temporal lobe lobectomy. Hippocampus 2019;29:63–7. 10.1002/hipo.22998.

[29] Kirschen RM, Leaver AM. Hearing function moderates age-related changes in brain morphometry in the HCP Aging cohort. BioRxiv Prepr Serv Biol 2024:2024.04.22.590589. 10.1101/2024.04.22.590589.

[30] Sarant JZ, Harris DC, Busby PA, Fowler C, Fripp J, Masters CL, et al. No Influence of Age-Related Hearing Loss on Brain Amyloid-β. J Alzheimers Dis JAD 2022;85:359–67. 10.3233/JAD-215121.

[31] Lad M, Taylor J-P, Griffiths TD, Alzheimer’s Disease Neuroimaging Initiative. Subjective hearing loss is not associated with an increased risk of Alzheimer’s disease dementia. Heliyon 2024;10:e30423. 10.1016/j.heliyon.2024.e30423.

[32] Lin FR, Pike JR, Albert MS, Arnold M, Burgard S, Chisolm T, et al. Hearing intervention versus health education control to reduce cognitive decline in older adults with hearing loss in the USA (ACHIEVE): a multicentre, randomised controlled trial. The Lancet 2023;402:786–97. 10.1016/S0140-6736(23)01406-X.

[33] Hansson O. Biomarkers for neurodegenerative diseases. Nat Med 2021;27:954–63. 10.1038/s41591-021-01382-x.

[34] Lad M, Taylor J-P, Griffiths T. Reliable Online Auditory Cognitive Testing: An observational study 2024:2024.09.17.24313794. 10.1101/2024.09.17.24313794.

[35] Bakkour A, Morris JC, Wolk DA, Dickerson BC. The effects of aging and Alzheimer’s disease on cerebral cortical anatomy: Specificity and differential relationships with cognition. NeuroImage 2013;76:332–44. 10.1016/j.neuroimage.2013.02.059.

